# Resident physician perspectives on ambient AI scribing in academic family medicine

**DOI:** 10.1101/2025.05.29.25328559

**Authors:** Himani Dhar, Angela Coderre-Ball, Akshay Rajaram

**Affiliations:** Department of Family Medicine, Queen’s University, Kingston, Ontario, Canada; Department of Emergency Medicine, Queen’s University, Kingston, Ontario, Canada

**Keywords:** Family practice/general practice/primary care, Medical students and residents, Medical education, ambient scribing

## Abstract

While ambient artificial intelligence (AI) scribes have been received positively by primary care physicians, the perceptions of resident physicians are not yet unclear. We conducted a qualitative study involving focus groups with first and second-year family resident physicians from a single urban academic family health team to gauge their understanding of ambient AI scribing and their perceptions of its potential impact on patient care. Seven resident physicians participated in two focus groups. Sessions were audio recorded and transcribed verbatim, then analyzed inductively to identify themes. We categorized the findings into five themes: 1) understanding of and exposure to AI, 2) perceived impact of ambient AI scribing on the practice of family medicine, 3) perceived impact on the cognitive load of charting, 4) performance and accuracy of ambient AI scribes, and 5) implications for adoption. Residents in this study reported minimal exposure to AI and concerns regarding the impacts of ambient AI scribing on the documentation process and quality of notes. Future research should explore the potential effects of ambient scribes on resident documentation prior to testing in practice.

## INTRODUCTION

Up to 25% of a primary care physician’s workday is spent on documentation of clinical notes^1,2^ and this increased administrative burden has been associated with decreased work satisfaction and increased burnout.^3-5^ Increasingly, evidence suggests ambient AI scribes reduce time spent on documentation,^6^ decrease workload and perceived burnout,^7^ and improve patient-physician rapport.^8,9^ While ambient AI scribes may be beneficial to attending physicians, less is known about their impact on resident physicians, who similarly spend significant time documenting in the EMR.^10-15^

Our primary objectives were to assess resident physicians’ understanding of AI and ambient AI scribing in healthcare and explore their perceptions of the potential impacts of this technology.

## METHODS

Our research adopts a qualitative grounded theory approach^16,17^ and took place at the Department of Family Medicine at Queen’s University, an academic family medicine teaching site in Kingston, Ontario between August 30 and November 7, 2023. We invited first-year resident physicians (PGY-1) and second-year resident physicians (PGY-2) to participate in a 30-to 45-minute focus group conducted over video-conferencing.^18^

One member of the authorship team (HD) facilitated all focus groups using a focus group developed by all authors (see Supplementary file 1). The facilitator was responsible for recording the discussion, asking questions, and clarifying participant questions related to the prompts. At the beginning of each focus group/interview a pre-recorded video demonstrating an ambient AI scribing program (see Supplementary file 2) was played for participants. Sessions were audio-recorded and transcribed verbatim by a hired transcriber.

Following the approach to thematic analysis by Braun and Clarke,^19^ two separate coders (HD, AR) independently performed manual line-by-line coding of the transcripts using NVIVO.^20^ They compared and discussed discrepancies to enable the development of a common codebook. The research team (HD, AR, ACB) met to review codes and build themes. The project received research ethics board approval from the Queen’s University Health Sciences Research Ethics Board (FMED-6877-23).

## RESULTS

### Demographics

Seven participants (three PGY-1, four PGY-2) participated in two focus groups (for PGY-1 and PGY-2 participants).

### Response Themes

#### Theme 1: Understanding and exposure to AI and AI applications in healthcare

There were differences in participants’ understanding of and exposure to AI in healthcare. Five of seven participants reported a basic understanding of AI and three of seven participants had a general understanding of the function of an ambient AI scribe (Table 1). Most participants reported minimal exposure to AI in healthcare. Only one participant reported exposure to ambient AI scribing software.

**Table 1.** Supporting quotations for themes 1 and 2.

| Theme | Summary | Quotations |
| --- | --- | --- |
| Understanding and exposure to AI and AI applications in healthcare | Residents described minimal experience with AI in healthcare, but many had a basic understanding of AI. | What I understand [an ambient scribe] to be is basically a microphone or some listening device inside the clinic room where it can overhear the conversation between a physician and the patient. Once you program the demographics, it can listen to the conversation and synthesize the main points. Then it can generate a note for that particular encounter.” PGY2 |
| Perceived impacts on patient care | Residents generally perceived that AI scribes had the potential to improve patient care by allowing them to focus on their patients during conversations, rather than typing notes. A few expressed concerns about speaking medical findings to the scribe with terminology that a patient might not understand. | “What’s nice about a clinical encounter is that it’s two humans sitting in a room and trying to figure out a set of problems... it has to fundamentally be that human interaction. And from a physician perspective that’s really rewarding. From an outcome perspective it’s particularly important and especially in family medicine where so much of it is based on the relationship.” PGY1<br>PGY-1: “I think personally it would really improve the physician-patient interaction because I feel like I’m always turned to the side looking at the computer while the patient is to a 90 degree to me. And I’m always typing, especially for mental health appointments. It’s always awkward if I have to type when they’re sad or tearful and it’s such an awkward interaction. Whereas if I could really give my like 100% to the discussion with the patient, I think it would really improve the quality of the interaction and the trust in the relationship that is built.”<br>PGY-2: “Or even if you’re describing your physical findings out loud, that may be unsettling to the patient because you’re talking about these ‘medical-ese’ and they don’t know what’s going on. |
|  |  | PGY-2: "... certainly with a mental health issue you need to give [patients] your whole attention. And that comes naturally, that's not something that gets forced. But having the additional scribe I think in the end would probably make things easier for the patient as well, since they can keep their thoughts together and kind of tell their own story rather than having gaps where you type and they're just waiting." |

#### Theme 2: Perceived impacts on patient care

Participants generally reported that looking at their screen to type disrupted the flow of the clinical encounter and felt this disruption negatively affected patient rapport, especially during conversations regarding mental health (Table 1). Participants reported a perceived benefit of ambient AI scribe use would be improved focus on patients (Table 1). However, some participants also described potentially detrimental effects, including speaking abnormal findings during physical examinations (Table 1).

#### Theme 3: Perceived impacts on the cognitive load of charting

Participants highlighted the potential decreased time spent on documentation when using an ambient AI scribe (Table 2) but emphasized the process of documenting as a valuable tool for learning and tracking clinically relevant details. Writing and reviewing notes was seen as a means for improving the structure of clinical encounters and served as a reflective process for learners as they considered conditions or ideas they may not have during the patient visit (Table 2).

**Table 2.** Supporting quotations for themes 3, 4 and 5.

| **Theme** | **Summary** | **Quotations** |
| --- | --- | --- |
| Perceived impacts on the cognitive load of charting | Residents described the perceived usefulness of writing notes for working through a diagnosis and remembering a patient. | "... I think we learn a lot as residents from writing notes. Whether it's retrospective, reflection on how the encounter went, versus these symptoms could actually mean this diagnosis that I didn't really remember and now I should follow up on it. Different things like that I think would be kind of lost if we used ambient scribes – that learning from writing and doing the grunt work early." PGY1<br>"... taking the note helps you remember things. I never thought about that but that's probably true for me because that's how I always studied: writing things down and typing things out." PGY2<br>"[Note-taking] lets me solidify that visit and lets me give myself a second viewpoint once I've stepped out of that environment. And that is something that I always depend on, just to make sure that I'm double checking myself and it gives me different balances and helps me organize my thoughts and get everything done appropriately. I think [the processes of reorganizing my thoughts and double-checking] would be lost because reading text that's already done for you is different than actually going through that mental process of reorganizing, rethinking, and |
|  |  | making it final, which I think is a very critical component of our learning as well as just being a physician.” PGY1 |
| Performance and accuracy of ambient scribes | Residents expressed concern for ambient scribes’ ability to understand non-linear conversations, variations in medical and lay jargon and potential biases. | “Lots of times, patients come in with two or three issues and then maybe you’re asking a question about the first issue and then move on to the second issue. And then you’re like – ‘oh wait I forgot to ask about this.’ How [the ambient scribe] puts it all together to organize multiple issues [is important because] depending on the patient or the appointment, you might be jumping a bit all over the place.” PGY2<br>“Patients complain of dizziness and you go through your differential. What’s going to get captured [by the ambient scribe]? Is that going to just cause more problems? And there’s lots of those words that can mean a lot of different things that would be kind of hard to capture without the context. I mean that’s always the difficult things with these kind of tools: how do you set for context?” PGY1<br>“Also there might be...medical jargon variation, or words that patients use that are different from the usual terminology, that’s used, especially in paediatric patients. I wonder how that would be interpreted by the scribe and if we need to clarify that during the conversation.” PGY1<br>“The overall problem that I worry about with AI is that it’s only as good as the data it learned on. And the phenomena of ‘pale male data’, there’s a lot more white people and there’s a lot more male built into all data sets. I do worry about the effect that it would have on biasing medical results.” PGY2 |
| Adoption of ambient scribes in family practice | Participants describe potential barriers to adoption in primary care including patient consent, data | “I think you’d have to get consent for recording. And then, if you’re getting consent, a certain amount of your patients are probably going to say – ‘oh that sounds a bit weird, I don’t like |
|  | security and medico-legal requirements of medical notes. | that.’ And then potentially you’ve broken down a little bit of trust. I’d probably have to think about the implications of that a little bit more, but that might be a deal breaker.” PGY1<br>“We’re already worried about EMRs getting hacked and whatnot. What about this technology? What is the chance of it becoming hacked, somebody else listening in on the conversation, losing the raw data, recordings that are very private to patients with health information? I don’t know how that would all work out. And where is this going to be stored? Is this going to be in a cloud? Where is the data actually going to be and how would it be protected?” PGY1<br>“I do not write down every single thing I talk about with a patient but this would. And then are you legally allowed to take things out of the transcript that are not relevant down the line? I’m not really sure how that would work out.” PGY2 |

Multiple participants commented that the cognitive process of writing notes differed between reviewing and proof-reading. Actively documenting encounters was thought to provide benefits in learning, consolidation, and reflection, while reviewing notes required primarily reading and correcting notes (Table 2). Participants also highlighted that ambient scribing could not include physicians’ thought processes.

Two PGY-1 participants commented on the format and standardization of ambiently scribed notes potentially varying from their own personal note-taking style and interfering with an individual physician’s preferred format of documentation. Participants felt that the process of standardization in this manner could result in the overinclusion of detail (Table 2).

#### Theme 4: Performance and accuracy of ambient scribes

Participants highlighted several concerns regarding the accuracy of ambient scribes in the context of speech differences across patients (accents, impairments or quieter voices), less structured encounters, and encounters with multiple people (e.g., healthcare workers, translators, and/or family members). Participants noted that in primary care, the discussion of multiple concerns in non-linear fashion may pose challenges for ambient AI scribes, and questioned how scribes would be able to assign the relevant details to each issue (Table 2).

Participants also raised concerns with the accuracy of transcriptions, including variations in terminology (e.g., “dizziness”), simple or colloquial terms, missed information or misinterpreted information, or simple terms used by paediatric patients. (Table 2). Additionally, one PGY-2 highlighted that a given ambient scribe’s accuracy would depend on the set of training data used and that these data could be biased (Table 2).

Multiple participants reflected on the medicolegal implications, including concerns around having incomplete or inaccurate documentation due to scribe use as well as the implications of making clinical decisions based on incomplete or inaccurate information. They also wondered about whether it would be legally defensible to remove information from the scribed documentation where unnecessary or irrelevant details were included.

#### Theme 5: Adoption of ambient scribes in family practice

Participants highlighted several barriers to the adoption of ambient AI scribing in family medicine, including policies and procedures around patient consent, training, and compatibility with different EMRs (Table 2).

Participants had mixed feelings regarding adoption of ambient AI scribes. They highlighted their experience adapting to multiple EMRs during residency and that having to learn an additional piece of software could be overwhelming. They felt adoption would take time and repeated use would be needed to gain comfort.

## DISCUSSION

This study explores the perspectives of resident physicians on the use of ambient AI scribes in an academic family medicine setting. Whereas residents acknowledged perceived benefits to ambient AI scribe use in terms of improved patient rapport, their limited exposure to this technology coupled with their concerns regarding its impact on documentation suggest the need for targeted education additional testing prior to its introduction in academic settings.

### The role of documentation in resident learning

Given significant charting burden,^10-15^ we were surprised to hear resident participants highlight the educational value of note writing and their preference for writing over editing in the context of ambient scribe deployment. Bowker et al. found that trainees used documentation as ways to support their clinical judgement, focus on patient issues, and identify gaps in knowledge and data collection.^21^ In this way, writing encounter notes is important to developing mental models about patients and ambient AI scribe use prior to the consolidation of these skills could be detrimental overall to resident learning.

### Documentation quality

Residents’ concerns regarding note quality resonate with a recent qualitative study that revealed attending physicians’ concerns with the accuracy, completeness and the formatting of notes following use of an ambient AI scribe.^7^ Given that residents receive limited formal learning opportunities around charting,^22^ developing a strong foundation through repeated practice of authoring notes is critical to identify potential errors or hallucinations in transcriptions.^23^

### Implications for adoption

These findings suggest that use of ambient AI scribing by residents in academic family medicine practices may not yet be ready for widespread deployment. To ensure residents continue to develop strong clinical problem-solving skills, including the management and synthesis of information about patients, residency programs could consider continuing resident notetaking alongside ambient AI scribing tools or limiting their use to senior years (e.g., transition to practice) with dedicated instruction beforehand around the medicolegal considerations.

## Limitations

Our sample size was small with participants reporting minimal exposure to AI, particularly in a practice setting. This observation is also likely due to the timing of this study, before widespread adoption of AI scribes in primary care in Canada. Furthermore, all participants were from one of four sites of an academic family medicine residency program. Accordingly, our findings may not be generalizable to other practice settings, including community and rural sites, longitudinal family medicine programs, or other specialties.

## CONCLUSIONS AND FUTURE WORK

We conducted a qualitative study to gather the perspectives of family medicine resident physicians on ambient AI scribing. While participants perceived benefits of ambient scribing in reducing documentation burden and improving physician-patient rapport, they raised concerns related to its educational impact and quality of documentation. In addition to addressing methodologic limitations, future work on ambient AI scribing should explore testing with family medicine residents in diverse settings to inform adoption.

## Data Availability

All data produced in the present study are not available.

## ACKNOWLEDGEMENTS

The authors wish to thank Dr. Noah Crampton of MutuoHealth for providing the ambient scribe demonstration used in this study.

## AUTHOR CONTRIBUTION STATEMENT

AR and ACB conceived the study. HD facilitated the focus groups with AR as an observer. Both coded the transcripts. ACB supervised and reviewed the codebook prior to analysis. All authors contributed equally to the writing and review of the manuscript. All writing was completed without the assistance of any AI or automated writing technologies. All authors agree with the content of the article and to its submission for publication.

## SUPPLEMENTARY MATERIAL

### S1. Focus Group Guide

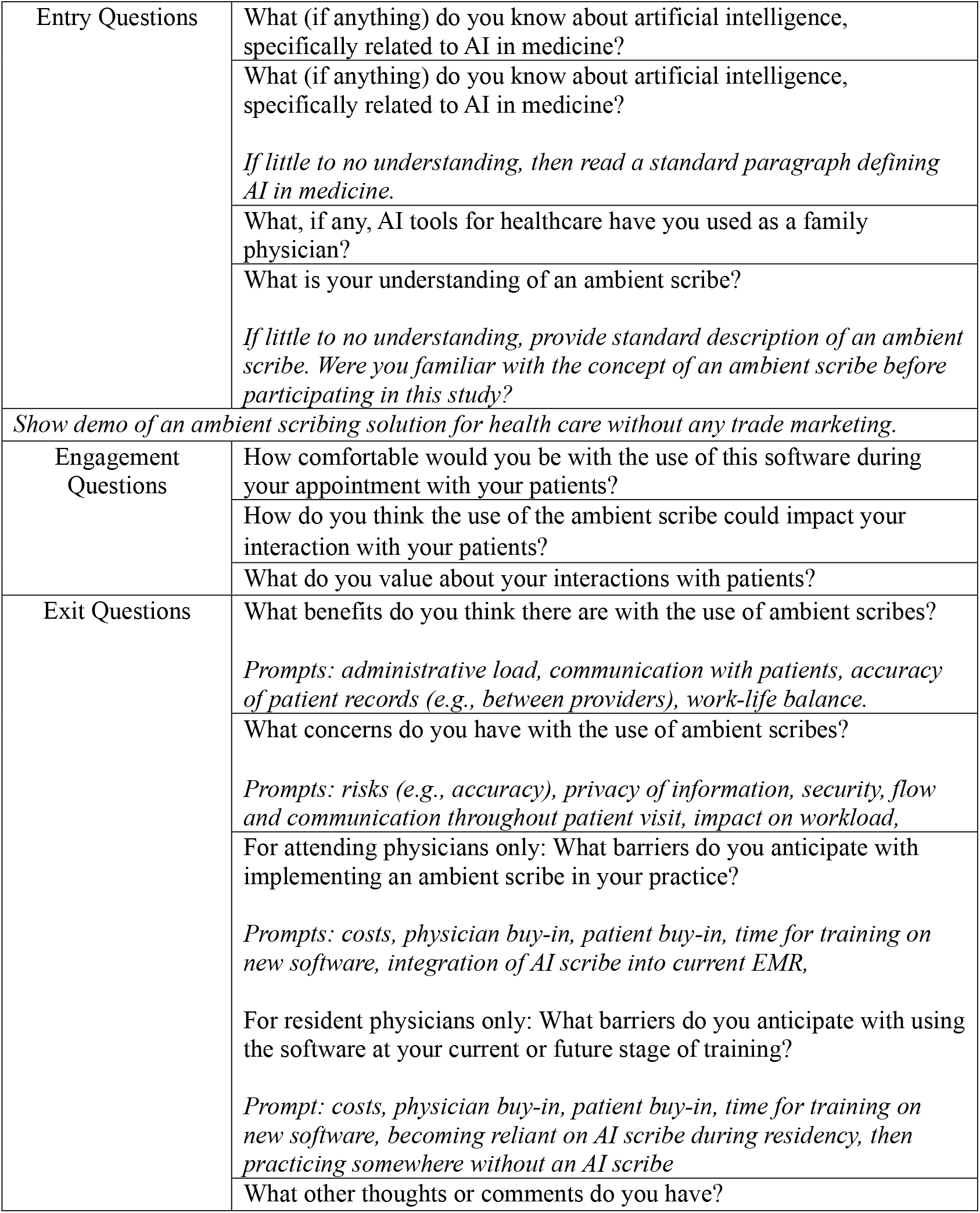

#### A.I. Definition and Summary

Artificial intelligence (or A.I. for short) is a technology that allows machines to perform tasks and make decisions similar to how humans do. AI uses algorithms, which are like step-by-step instructions, to process huge amounts of data and find patterns or solve problems. It can recognize images and understand speech, which can allow it to help with a variety of complex tasks. AI can also adapt and improve over time by learning from its experiences. An example of an A.I. software you may have heard of is Chat-GPT.

#### Ambient scribe Definition and Summary

An ambient scribe is an A.I. technology that helps people capture and record information in a digital format. They can take notes and transcribe spoken words into written text. They can be helpful in summarizing conversations into a standardized format. This technology is especially helpful for individuals who need to keep accurate records of meetings, lectures, or interviews. Ambient scribes can be used on computers, smartphones, or other devices and save time and effort by eliminating the need to manually write everything down.

### S2. Video demonstrating an ambient scribe

https://bit.ly/perspectives_supplementary2

